# Effectiveness of a family-led postnatal care model: A pre-post intervention pilot study in the Ada’a District, Ethiopia

**DOI:** 10.64898/2026.03.04.26347595

**Authors:** Gadise Bekele, Della Berhanu, Konjit Wolde, Dedefo Teshite, Lello Amdissa, Walelegn Worku, Pooja Sripad, Anne Hyre, Lisa Noguchi, Alemayehu worku

## Abstract

**Background:** Postnatal care is crucial for assessing and improving the health of both mothers and newborns, yet its coverage remains low in Ethiopia. Timely, high-quality postnatal care, especially within the first week after birth, is essential to reduce maternal and neonatal mortality. Family-led postnatal care is an innovative model for reaching postnatal mothers and newborns during the first week after birth. Leveraging self-care principles, mothers, with the support from family and guided by a checklist, perform daily postnatal health checks on themselves and their newborns. This study evaluated the effect of a family-led postnatal care intervention on coverage of postnatal checks within seven days of birth.

**Methods:** This study used pre- and post-intervention cross-sectional surveys in four health centers. Eligible postnatal mothers who gave birth in the study health centers were interviewed pre- (November 2022 to January 2023) and post-intervention (February to April 2023) using a structured questionnaire. Bivariate tests and descriptive analyses were used to assess changes in postnatal care coverage over time.

**Results:** Surveys were completed with a total of 119 mothers pre-intervention and 110 mothers post-intervention. In the pre-intervention period, 9% (11/119) of mothers and 11% (13/119) of newborns had a postnatal check between 24 and 72 hours after birth, whereas in the post-intervention period this increased to 96% (105/110) mothers and 96% (105/110) newborns (P<0.0001). A similar increase occurred in the proportion of mothers and newborns having postnatal checks between 73 hours and 7 days (3% vs. 96%, P< 0.0001). Compared to pre-intervention, a larger proportion of mothers detected a maternal danger sign during the post-intervention period (6.7% vs 18.2%, P<0.008).

**Conclusion:** Family-led postnatal care is a promising self-care model that may increase postnatal checks for mothers and newborns who would not otherwise have received care. Evaluating this model in other settings using a more rigorous design is recommended.

**Trial registration:** ClinicalTrials.gov (NCT05563974), first posted on 3 October 2022.

## Background

Postnatal care (PNC) is defined by the World Health Organization (WHO) as care given to the mother and her newborn immediately after birth and for the first 42 days after.(1) To ensure maternal and newborn wellbeing, WHO recommends that all postnatal mothers and newborns have at least four postnatal checks: within 24 hours after birth; between 24-72 hours; between 72 hours and seven days; and at 42 days(2). Despite its importance, the postnatal period is the most neglected along the continuum of maternal and newborn care.(3) Half (49%) of maternal deaths occur on the first day after childbirth and 25% of deaths occur between days two and seven.(4) Similarly, half of all neonatal deaths occur in the first day of life, and nearly three quarters occur within the first week of life.(5,6) Ensuring regular high-quality PNC visits, particularly within the first week after birth, is paramount to reduce maternal and neonatal mortality.(4,7–11)

The federal government of Ethiopia has targets to increase coverage of early PNC within two days from 34% to 76%, decrease the maternal mortality ratio from 401 to 279 deaths per 100,000 live births, and reduce the neonatal mortality rate from 33 to 21 deaths per 1,000 live births by 2017 Ethiopian fiscal year (2024/25).(6) According to the 2016 Ethiopian Demographic and Health Survey (EDHS),, only 17% of mothers and 14% of newborns receive at least one PNC service from a health institution in the first 48 hours after birth.(12) Per the national Community Based Newborn Care guidelines, health extension workers should conduct home visits for all postnatal mothers in their communities.(13,14) However, these home visits are not occurring due to health extension workers’ heavy workload, poor supervision, and lack of knowledge that a birth has occurred; inadequate drug supply; distance to homes; and topography of their catchment areas.(14–16) At the same time, families generally do not seek PNC at health facilities due to their perceptions of being healthy, lack of awareness of the service, negative attitudes of health care providers and lack of information on the importance of PNC.(17)

Human-centered design (HCD) activities were conducted to better understand the challenges with timely PNC use in the local context. Findings revealed that mothers and newborns were expected to stay home during the first 42 days after birth and particularly the first 10 days. In the postnatal period, mothers most trusted their husbands and immediate family members, religious leaders, and neighbors when seeking care/advice or making decisions. Postnatal mothers also felt they were expected to “prove” themselves as good mothers, and feared not knowing how to care for their newborn, or how to recognize problems. Postnatal mothers and their families tended to seek care from health facilities as a last resort, when they were in dire need.(18)

Based on these insights, the family-led postnatal care (FPNC) intervention was designed as an innovative model for reaching postnatal mothers and newborns with key PNC services during the first week of life that leverages self-care principles.

The WHO defines self-care as “the ability of individuals, families and communities to promote health, prevent disease, maintain health and cope with illness and disability with or without the support of a health worker”(19). Self-care interventions include evidence-based, high-quality drugs, devices, diagnostics and/or digital interventions that can be provided fully or partially outside formal health services and be used with or without a health worker to support self-care.

The Ethiopian Ministry of Health has developed a major strategy and guidelines to support the introduction and implementation of self-care interventions for improving maternal and newborn health.(6). Experience with self-care interventions, implemented with support from the health system during the Corona virus disease of 2019 ( COVID-19) pandemic, generated ample evidence on their value for both general and maternal health care.(19–23). These studies have shown promising results indicating the use of innovative self-care interventions that may impact maternal and neonatal morbidity and mortality.

The objective of the current study was to evaluate the effect of family-led postnatal care intervention on coverage of postnatal check within seven days of childbirth for both the mothers and newborns.

## Methods

### Study setting

Primary level healthcare in Ethiopia is provided by a health center (where most facility-based deliveries take place) and approximately five catchment health posts, where two health extension workers provide curative and preventative services. The study was conducted in four health centers and their catchment *kebeles* in Ada’a district of Oromia Region, Ethiopia. The population of Ada’a is 204,847 with 7,109 annual births. At the time of the study, Ada’a district had 5 health centers, 22 *kebeles*, 22 health posts and 37 health extension workers during the study period. One of the five health centers, was excluded as it was part of the conceptualization, design, and testing of the FPNC intervention.

### Study design

This study used quantitative pre- and post-intervention cross sectional surveys. The pre-intervention survey was conducted from November 14, 2022 to Jan 22, 2023, and the post-intervention survey from February 6 to April 9, 2023. The intervention was implemented after the pre-intervention data collection was completed.

### Study participants

Mothers who delivered at the four participating health centers were approached to participate in the study. Mothers who were within three days of birth during discharge from the health center, intended to remain within the catchment area of the health center for the first week after birth, had family members who were willing to participate in the study, and were willing and able to provide consent were included in the study.

#### Program Description

The FPNC intervention consisted of several components (Figure 1). At the health center, a pre-discharge postnatal assessment and counseling is conducted by the health center staff, guided by a discharge counseling poster. A health care provider invites family members into the postnatal room to observe how they perform PNC tasks for the mother and newborn using visual checklist. The health center staff checks for: how the mother feels; severe headache; vaginal bleeding; breast condition; blood pressure; temperature; and swelling in the face and leg. The pelvic examination is done privately without the family members. The health center staff checks: if the newborn appears to be feeling well; is breastfeeding well; and the newborn’s skin color, umbilical cord conditions, breathing pattern, temperature, and vaccination status. The checklist is completed by the staff during the pre-discharge checks.

**Figure 1:**
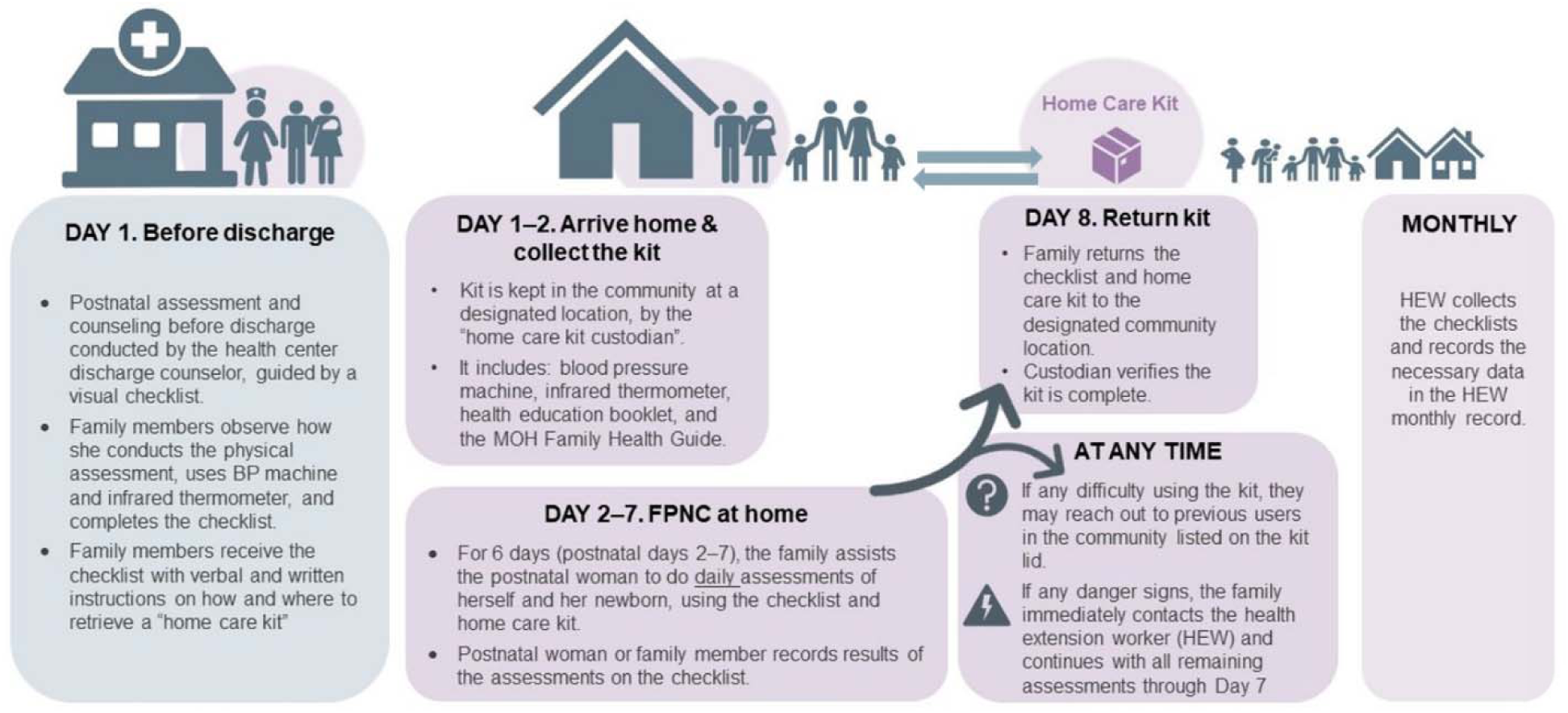
Summary of the family-led postnatal care intervention, Ada’a district, Oromia region of Ethiopia, 2023

Upon discharge, the health center staff gives the checklist to the family with verbal and written instructions on how and where to retrieve a “home care kit” (HCK) in their community. The HCK includes a blood pressure machine, infrared thermometer, a pictorial health education booklet designed for both literate and non-literate users, and a checklist for guiding daily self-care tasks and recording results.

Three homecare kits are placed in one *kebele* (i.e., the lowest administrative unit in Ethiopia) consisting of approximately 10,000 households. The kits are placed in the homes of community members or leaders identified by health extension workers who serve as homecare kit “custodians”. After arriving home, a family member retrieves the HCK from the designated custodian, who records the check-out date and contact information of the family member in a HCK register. For six days (postnatal days 2–7), the postnatal mother, with the assistance from family members, conducts daily assessments of herself and her newborn, using the checklist and contents of the HCK. If a family has difficulty using the devices, they are instructed to reach out to the HCK custodians, health extension workers, or other families who have previously used the HCK in the community. A log on the lid of the HCK indicates who else in the community has used the HCK along with their contact information. An additional file shows the discharge counseling poster, visual checklist, HCK, education booklet and HCK register in more detail [see Additional file 1].

The postnatal mother or family member records the results of the assessments on the checklist. If the assessment reveals any danger signs, the checklist instructs the family to immediately contact a health extension worker and to continue with the remaining assessments up to the seventh day. Around the eighth day after delivery, the family returns the HCK and completed checklist to the custodian, who verifies the availability and functionality of the equipment and records the HCK return in the register. The custodian asks the family for permission to list their contact information on the HCK log in case other families need guidance on how to use the devices. If the family member grants permission, the information is recorded on the HCK log. At least monthly, the health extension worker from the closest health post collects the completed checklists and records the assessments in the Integrated Maternal Newborn Child Health Card.

### Sample size and sampling

The sample size was calculated using the 2016 EDHS coverage of PNC within 24 hours of 17%.(12) With a desired increase in PNC coverage to 45% due to the intervention, a 5% level of significance, 80% power, a design effect of 2.0, and a non-response rate of 10%, the minimum sample size required to detect the assumed difference was 109 postnatal mothers at both pre-intervention and post-intervention each, for a total of 218 postnatal mothers. Postnatal mothers who delivered at the study health centers were offered participation sequentially until the sample size was fulfilled.

### Data collection procedures

Training was provided to field data collectors and supervisors on the study objectives, procedures, and ethics. A detailed survey manual with extensive standard operating procedures was prepared and used in the training, piloting, and survey itself. The training also included proper use of tablets for data entry at the point of collection.

Health center staff who were involved in the postnatal discharge process at the four selected study health centers screened mothers for participation in the FPNC study. Before discharge from the study health center, the health center staff asked screened mothers if they were interested in speaking with an FPNC study staff to learn more about the study. If the mother expressed interest, she was referred to the FPNC study staff. The study staff member who was on site approached all mothers and their accompanying family members to assess further eligibility, explain the study procedures, answer any of their questions, and obtain informed consent. If eligible and consented, study staff obtained contact information from the mother/family member. Study staff communicated the contact information for the local study enumerator. Data collectors assigned to the respective *kebeles* interviewed these mothers on the eighth postnatal day at their home.

### Outcome measurement

Postnatal mothers were interviewed using a structured questionnaire to collect information on basic socio-demographic data (age, educational status, and marital status). In addition, the following information on the primary and secondary outcomes was collected.

Primary outcomes:

- Proportion of mothers and newborns who had a postnatal check within 24 hours, 24–72 hours, and 73 hours–seven days;
- Proportion of mothers and newborns who assessed key PNC components within 24 hours, 24–72 hours, and 73 hours–seven days (based on self-report in the pre-intervention and from checklists provided to families in the post-intervention); and
- Average number of days any PNC assessed (mothers and newborns).

Secondary outcomes:

- Proportion of postnatal mothers who identified having a danger sign;
- Proportion of newborns with a danger sign identified; and
- Among those with an identified danger sign, the proportion of mothers and newborns who sought care from a health provider (disaggregated by type of provider and type of facility)

### Data management

Data were collected electronically using SurveyCTO designed tablets. Data from the field were synchronized daily to the central cloud server. There were daily checks on completeness and consistency of the collected data. The electronic data capturing system had built-in logic, range, and skip-patterns to limit data inaccuracies. Data were cleaned before the main analysis.

### Data analysis

Among all participating mothers, the proportion of mothers who had a postnatal check at each of the three time points (within 24 hours, 24–72 hours, and 73 hours-seven days) was computed and compared between the pre- and post-intervention periods using a chi-square test. This was repeated for newborns. Further descriptive analyses of differences in the recognition of and care-seeking for any postnatal maternal or neonatal danger signs and symptoms before and after the intervention was conducted to demonstrate FPNC secondary effects. STATA version 15 software was used to analyze the data.(24)

## Results

In the pre-intervention survey, a total of 144 mothers were screened, of which 119 met eligibility criteria and consented to participate in the study. After the introduction of FPNC (post-intervention phase) 133 mothers were screened, of which 115 met the eligibility criteria and 110 consented to participate in the study and took the checklist home. Of the 110 mothers who took the checklist home, 105 had family members collected the homecare kit (Figure 2).

**Figure 2:**
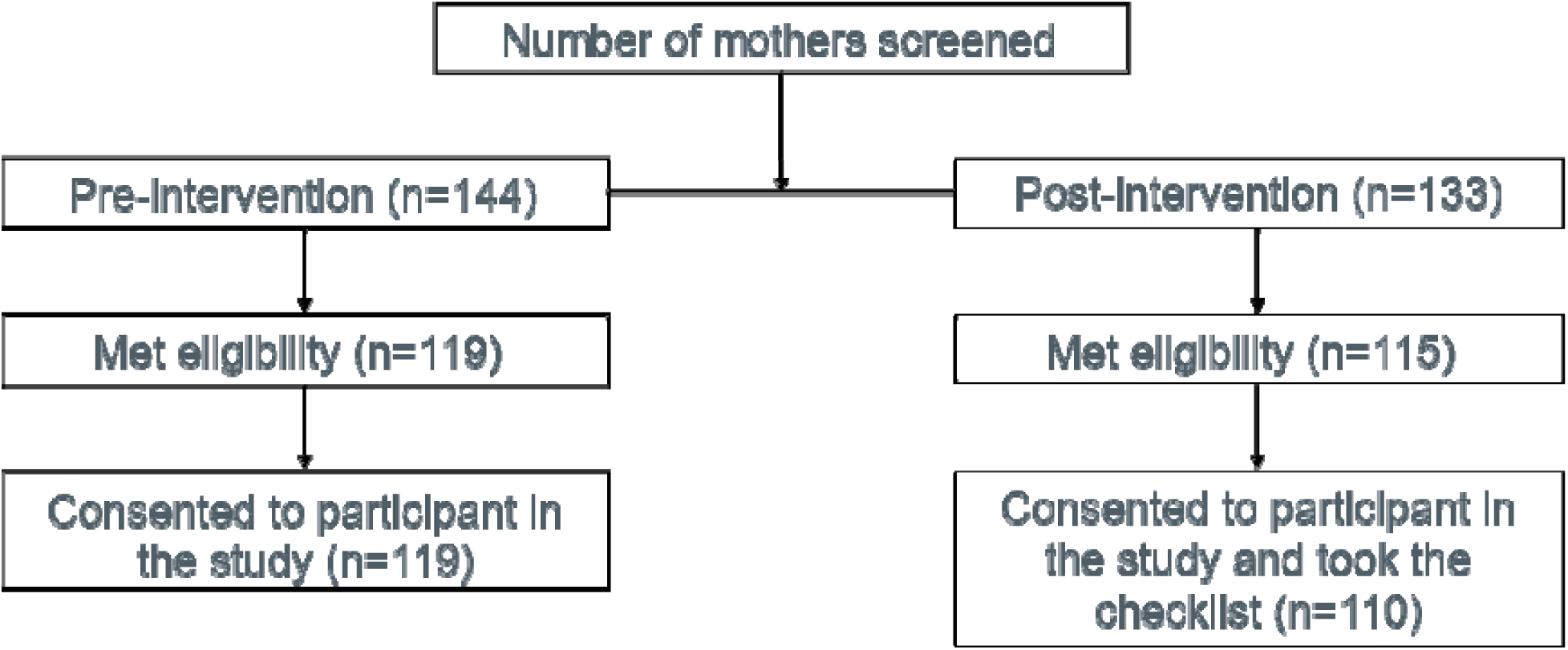
Participants flow diagram, Ada’a district, Oromia, Ethiopia, 2023

During both the survey periods, approximately 40% were between 18-24 years of age and around 45% were between 25-34 years of age (Table 1). Distribution of the educational level, religion, and martial status for mothers were comparable between the two time periods. Approximately a quarter of mothers had no formal education, and half had a primary level education. Ethiopian Orthodox was the predominant religion (>85%). More than 90% of respondents in both the pre- and post-intervention surveys were either currently married or cohabitating.

**Table 1:** Background characteristics of mothers, Ada’a district, Oromia, Ethiopia.

|  | **Pre-intervention** | **Post-intervention** |
| --- | --- | --- |
|  | **N= 119** | **N=110** |
|  | **n (%)** | **n (%)** |
| **Age in years** |  |  |
| 15-17 | 3 (3) | 1 (1) |
| 18 - 24 | 48 (40) | 44 (40) |
| 25- 34 | 55 (46) | 49 (45) |
| ≥ 35 | 13 (11) | 16 (15) |
| **Education level** |  |  |
| None | 29 (24) | 26 (23) |
| Primary | 64 (54) | 58 (53) |
| Secondary | 24 (20) | 25 (23) |
| University | 2 (2) | 1 (1) |
| **Religion** |  |  |
| Orthodox | 102 (86.7) | 96 (87) |
| Protestant | 16 (13) | 12 (11) |
| Muslim | 1 (1) | 2 (2) |
| **Marital status** |  |  |
| Never married | 6 (5) | 5 (5) |
| Currently married | 89 (75) | 87 (79) |
| Separated | 4 (3) | 1 (1) |
| Refused to answer | 1 (1) | 1 (1) |
| Cohabiting | 19 (16) | 16 (14) |

### Coverage of postnatal checks within 24 hours, 24 – 72 hours, and 73 hours - seven days

As shown in Table 2, in the first 24 hours after birth, 97% of the mothers and newborns had a postnatal check in the pre-intervention survey, while 100% of the mothers and newborns had a postnatal check in the post-intervention period (P = 0.053). In the pre-intervention survey, less than 11% of mothers and newborns had a check between 24 and 72 hours, and 3% had a check between 73 hours and seven days postpartum. In the post-intervention period, 96% of mothers and newborns had a check between 24 and 72 hours and between 73 hours and seven days postpartum (P <0.0001).

**Table 2:** Postnatal checks for mothers and newborns in the first week, Ada’a district, Oromia, Ethiopia.

| Timing of postnatal checks | Mothers |  |  | Newborns |  |  |
| --- | --- | --- | --- | --- | --- | --- |
|  | Pre-intervention<br>N=119 | Post-intervention<br>N=110 | P value | Pre-intervention<br>N=119 | Post-intervention<br>N=110 | P value |
|  | n (%) | n (%) |  | n (%) | n (%) |  |
| Within 24 hours | 115 (97) | 110 (100) | 0.053 | 115 (97) | 110 (100) | 0.053 |
| 24-72 hours | 11 (9) | 105 (96) | <0.0001 | 13 (11) | 105 (96) | <0.0001 |
| 73 hours- | 3 (3) | 105 (96) | <0.0001 | 3 (3) | 105 (96) | <0.0001 |
| seven days |  |  |  |  |  |  |

### Assessment of postnatal signs and symptoms

During the pre-intervention period, in the first 24 hours after birth, mothers reported being checked for general headache (57%), breast condition (53%), bleeding (48%), blood pressure (42%), temperature (39%), and face and leg swelling (9%). Post-intervention, checklists revealed that mothers and families conducted self-checks at home for postnatal danger signs and symptoms, which resulted in nearly all (>95%) mothers having the above-mentioned checks. Pre-intervention, from 24 hours to 72 hours and 73 hours to seven days, less than 10% of mothers had these signs and symptoms checked, while post-intervention over 95% had all study-specified signs and symptoms checked during the specified time period (Figure 3). This change is statistically significant (p<0.0001).

**Figure 3:**
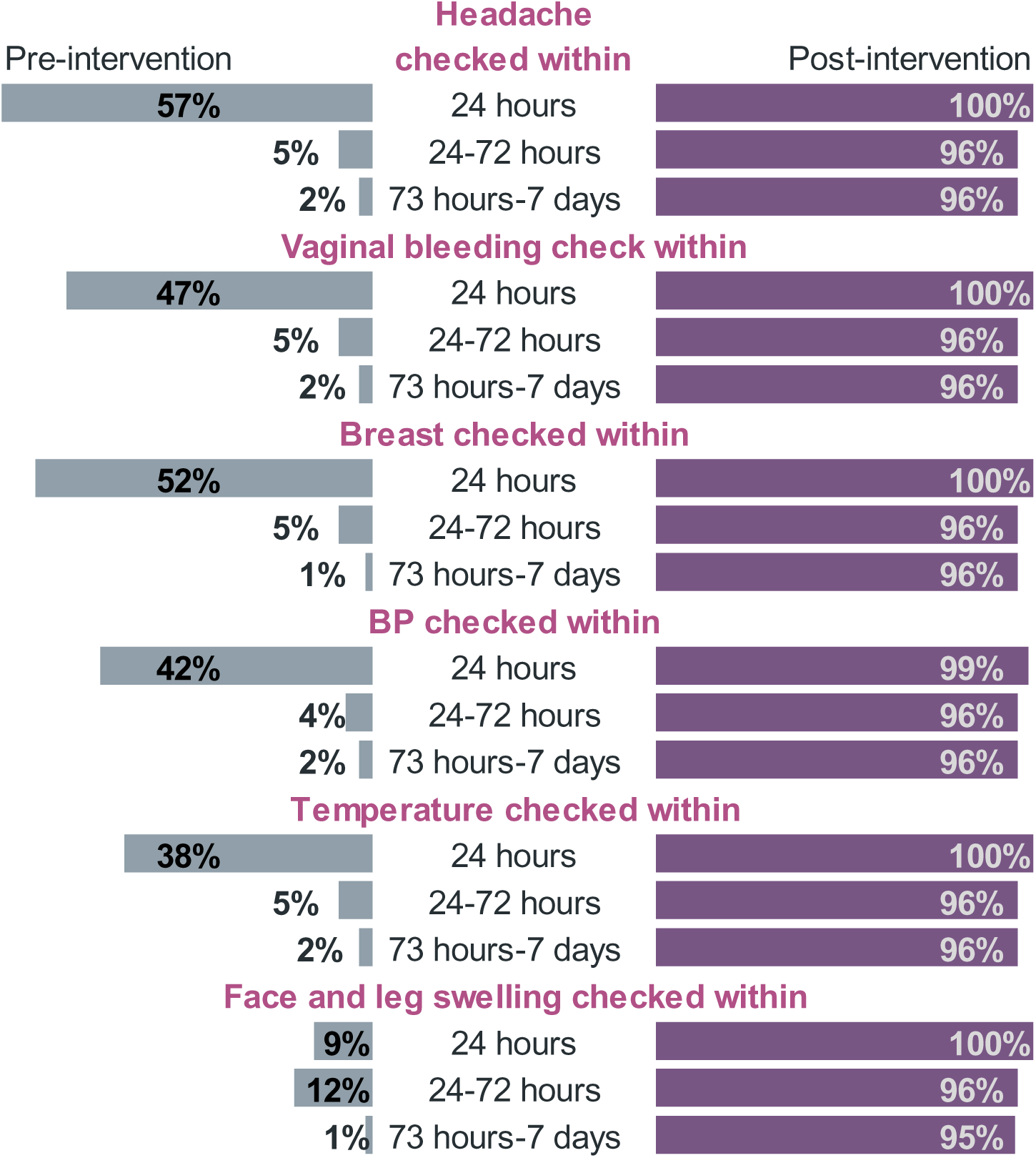
Maternal signs and symptoms checked during the first week after birth, Ada’a district, Oromia, Ethiopia

For newborns, during the pre-intervention period, in the first 24 hours after birth, mothers reported checks for breastfeeding (79%), umbilical cord (79%), general wellbeing (68%), breathing pattern (47%), temperature (36%), and skin color (25%). During the post-intervention period, almost all (>95%) newborns had all the above-mentioned checks in the first 24 hours after birth. Pre-intervention, from 24 hours to 7 days, less than 10% of newborns had these signs and symptoms checked, while post-intervention over 95% (p<0.0001) had all of the signs and symptoms checked in the same time period (Figure 4).

**Figure 4:**
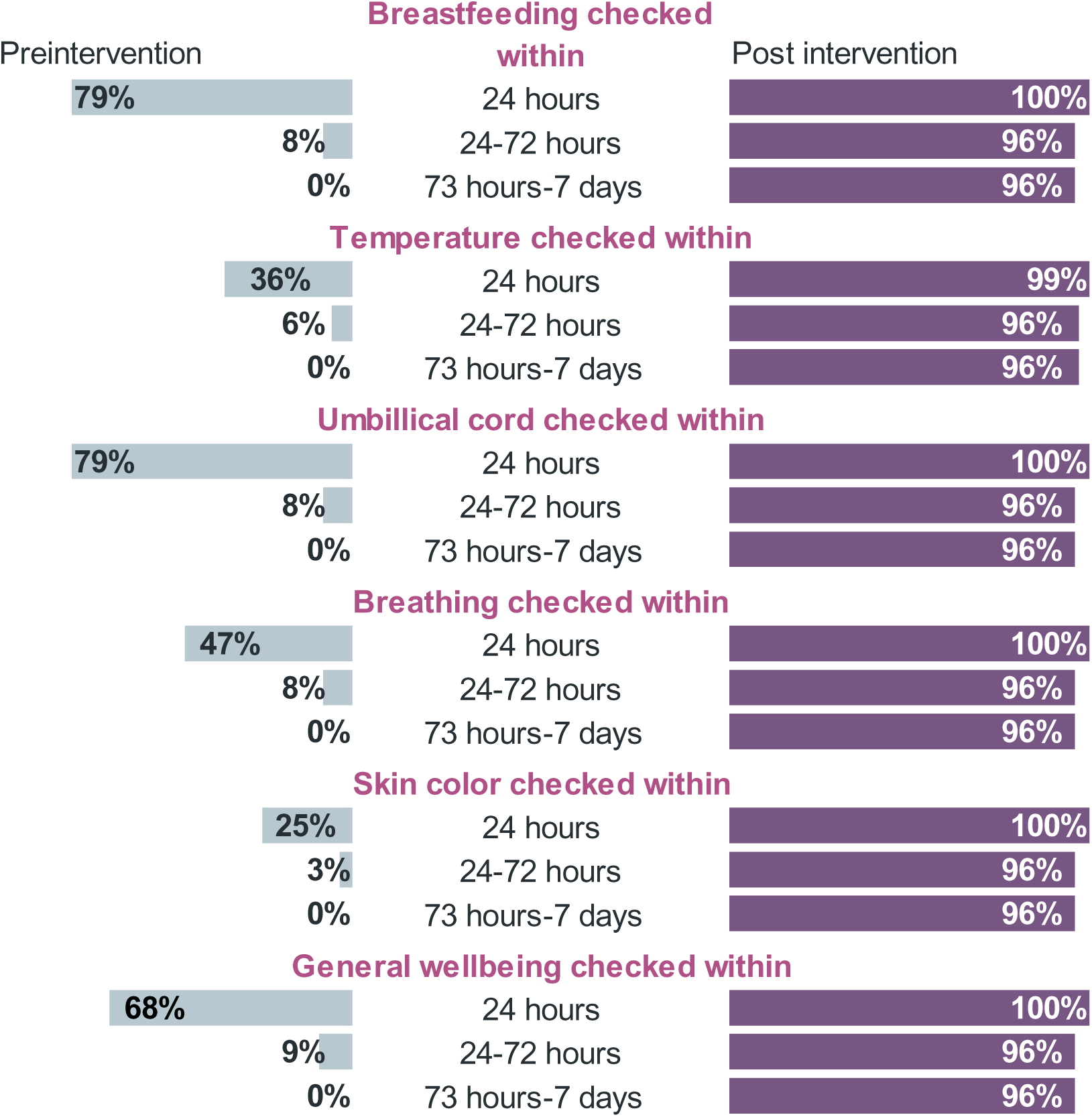
Newborn signs and symptoms checked during the first week after birth, Ada’a district, Oromia, Ethiopia

### Detecting and seeking care for danger signs

As shown in Table 4, pre-intervention, 7% of mothers detected a danger sign, while post-intervention, 18% detected a danger sign (p = 0.008). Among those who detected a danger sign, 63% in the pre-intervention period sought care (from health centers), while in the post-intervention period, 80% sought care (i.e., 65% from health centers, 15% health posts) (p = 0.4). During pre-intervention period, mothers detected headache (3%), bleeding (2%), breast conditions (2%), blood pressure problem (2%) and temperature problem (1%). Post-intervention, beyond identifying high blood pressure problem (11%), headache (5%), breast conditions (5%), and vaginal bleeding (4%), mothers additionally detected temperature problem (3%) and swollen face and leg (2%) as danger signs and symptoms (data not shown). An additional file shows the data on detected danger sign of the mothers in more detail [see Additional file 2].

Among newborns, there was no significant difference in the detection of danger signs between study periods (pre-intervention 4% vs. post-intervention 3%; p=0.5). Among families who detected a danger sign, 80% sought care (from health centers) pre-intervention, while 100% sought care (i.e., 67% from health center, 33% from health post) post-intervention (p = 0.6) (Table 4). Newborn signs and symptoms detected pre-intervention were breastfeeding problems (3%), umbilical cord conditions (3%), temperature problems (1%), and problems with breathing pattern (2%). In the post-intervention period, umbilical cord conditions (2%) and skin problems (1%) were the danger signs detected (data not shown). An additional file shows the data on detected danger sign of the newborn in more detail [see Additional file 2].

**Table 3:**
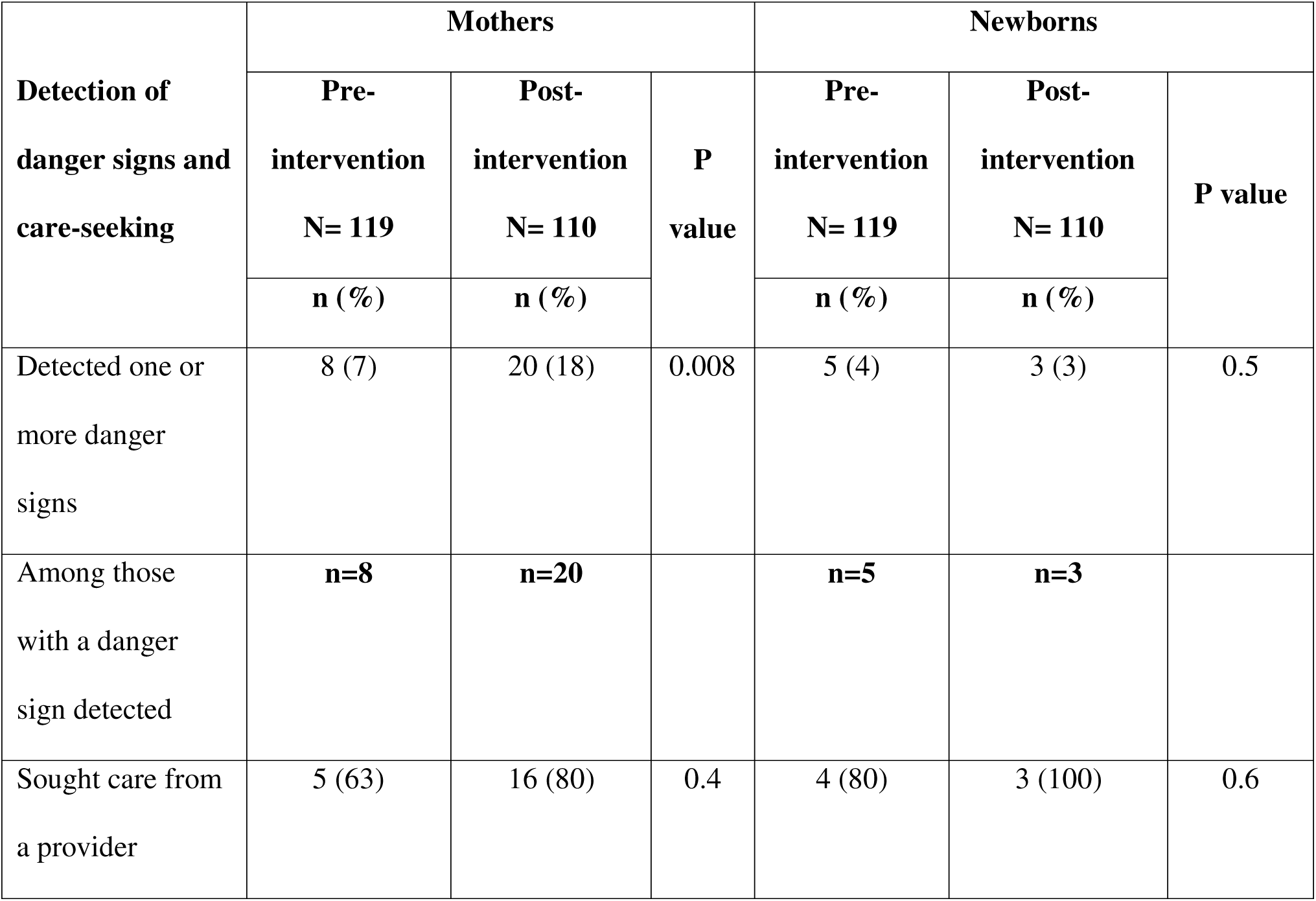
Danger sign detection and care-seeking during the first week after birth, Ada’a district, Oromia, Ethiopia.

Compared to pre-intervention, the post-intervention survey showed an increase in the proportion of mothers who reported having knowledge on how to detect a problem in themselves (83% vs. 97%, p<0.001) and their newborns (92% vs. 100% p=0.002). Similarly, an increase in knowledge was noted on what actions to take for a problem detected in themselves (88% vs. 97%, p=0.009). No change was seen for actions to take for a newborn problem (98% vs. 99%, p=0.67) (Table 5).

**Table 4:** Mothers’ perception and knowledge of health problem detection, Ada’a district, Oromia, Ethiopia.

|  | Pre-intervention<br>N=119 | Post-intervention<br>N=110 | p-value |
| --- | --- | --- | --- |
|  | n (%) | n (%) |  |
| Mother knows: |  |  |  |
| How to detect she has a problem | 99 (83) | 107 (97) | <0.001 |
| What actions she should take if there is a maternal problem | 105 (88) | 107 (97) | 0.009 |
| How to detect that her newborn has a problem | 110 (92) | 110 (100) | 0.002 |
| What actions she should take if there is a newborn problem | 117 (98) | 109 (99) | 0.67 |

## Discussion

This study aimed to assess if a family-led self-care model for postnatal care could increase the coverage of postnatal checks during the first week after birth. Nearly all participant mothers during the pre- and post-intervention periods had postnatal checks in the 24 hours after birth. Pre-intervention, one in ten mothers and newborns had a postnatal check between 24 and 72 hours after birth, whereas during the post-intervention period, nine in ten mothers and newborns had a postnatal check. A similar increase was noted in the proportion of mothers and newborns having postnatal checks between 73 hours and seven days. Results from this study suggest that mothers supported by their families, doing and documenting their home-based checks led to the increased number of postnatal checks in the post-intervention phase. Compared to pre-intervention, a larger proportion of mothers detected a maternal danger sign during the post-intervention period (7% vs. 18%) and sought care (63% vs. 80%). In contrast, very few families detected a newborn danger sign in either pre- or post- (4% vs. 3%) intervention periods, and nearly all sought care (80% vs. 100%).

This study has several notable strengths, particularly the use of a locally co-designed and led intervention created to be compatible with community cultural practices and beliefs, which was evaluated in a real-world setting. The intervention leveraged multiple key priorities of local government with regard to improving coverage of PNC and increasing access to self-care interventions. However, several limitations should be noted. Several key assessments for mothers were not part of the FPNC package, including heart rate, depression and anxiety. First, this pilot study did not randomize participants to intervention versus control. Use of a pre-/post-design creates the possibility that effects were due to period-related influences other than the study intervention. However, the pre- and post-intervention periods were relatively short (eight weeks each with a one-month implementation period in between) and thus unlikely for other interventions or time-based factors to have contributed to the observed changes in study measures. In the pre-intervention period, we used self-report to collect data on postnatal checks which could be prone to recall bias. However, the data collection on the 8th day after delivery aimed to minimize recall bias. The study’s reliance on self-report for some measures, including aspects of checklist completion, may have been impacted by social desirability bias. The number of participants enrolled was relatively small, though the study is powered; thus, generalizability is likely to be limited.

Completion of postnatal checks in the first 24 hours was high for mothers and newborns during the pre- and post-intervention periods, with almost all reporting completion of a check. This contrasts with data from the EDHS 2016, which showed that less than two in ten mothers and newborns received a check in the first 48 hours.(12) The difference in findings between the two studies may be because our data collection was limited to mothers having facility deliveries, while EDHS included both home and facility births. The FPNC intervention did not appear to impact completion of postnatal checks in the first 24 hours, likely because the model is initiated at the facility level where most if not all mothers who give birth have at least one postnatal check prior to discharge.

Despite high coverage of postnatal checks in the first 24 hours in both pre and post-intervention periods, the quality of the checks provided by the health center staff improved in the post-intervention period. Per the country guidelines, PNC checks for mothers should assess signs and symptoms of bleeding, pre-eclampsia, and sepsis, amongst others. Pre-intervention, only four in ten mothers were checked for bleeding, blood pressure, and temperature. Although low, this proportion was much higher than reported in another study.(16) In contrast, in the post-intervention period almost all mothers had their uterine bleeding, blood pressure, and temperature checked.

Similarly, pre-intervention in the first 24 hours, eight in 10 newborns had their cord checked and were assessed for breastfeeding, and one in three had their temperature checked. This was much higher than what was reported in the Mini-EDHS 2019 for cord check (26%) and breastfeeding assessment (34%), while temperature measurement was comparable (26%).(25) After the FPNC intervention was implemented, almost all newborns had all recommended components of the postnatal checks completed. The increased comprehensiveness in the post-intervention period can likely be attributed to the use of checklists by the health center staff during discharge. Hence, it implies that systematic and improved discharge counselling process for postnatal mother and family and integrating a checklist as tracking tool may have increased the quality and comprehensiveness of postnatal care. Moreover, the involvement of the family members in the discharge counselling likely contributed to better follow up of the postnatal mother and newborn.

Per national guidelines, health extension workers are expected to provide home visits on the third and seventh day. Yet, pre-intervention, very few mothers and newborns received the recommended number of postnatal checks between 24 hours and 7 days. These findings are similar to those of a study that showed about one in ten mothers who delivered in a facility received a postnatal check between 24 and seven days after delivery.(16) Post-intervention, 96% of families retrieved the home care kit and almost all mothers and newborns received postnatal checks between 24 hours and seven days. This implies that the home care kit, checklist and family engagement act as resources and reminders for the mothers and families to conduct daily health checks that detect key maternal and neonatal danger signs.

In this study, pre-intervention 7% of mothers detected a danger sign of which 63% sought care from a provider. Our results in the pre-intervention period differ from findings from a study in Kenya that found only 28% of postpartum mothers who detected danger sign sought care.(26) After the intervention, the proportion of mothers who detected a danger sign increased (18%) and a majority (80%) sought care. This is similar to reports from previous studies where mothers assessed themselves based on a text message sent to them in the postpartum period.(26,27) Use of the text messages in these studies and engagement of families served as reminders to conduct health checks and consequently seek care when detecting danger signs. These findings suggest that it is possible to directly engage mothers and influence care-seeking using this innovative self-care model.

For newborns in both the pre- and post-intervention surveys, very few families detected a danger sign, and almost all sought care. The number of families in this study who detected a danger sign was lower in both time periods as compared to the aforementioned Kenya study.(26) This may be attributed to the difference in the data collection period, where the other study collected data on danger signs in a two-month period after birth as compared to the one-week period used in this study and different study areas. For mothers and newborns, pre-intervention, those who sought care did so from the health center while post intervention, some sought care from health posts as well.

## Conclusions

In conclusion, FPNC is a promising, self-care intervention that has potential to increase coverage and quality of postnatal care for mothers and newborns who would not otherwise have received care. Evaluating this model in other settings with a more rigorous design is recommended. Future studies can also investigate if this model also works for mothers who are unable to deliver in health facilities.

## List of abbreviations

## Declarations

## Ethical considerations

Ethical approval was obtained from the Johns Hopkins Bloomberg School of Public Health Institutional Review Board (IRB No. 21096) and the Addis Continental Institute of Public Health Institutional Ethical Review Board (IRB No. 0029). Participation in the FPNC study was entirely voluntary. All postnatal mothers identified in the study who met the inclusion criteria were offered enrollment. In the post-intervention period, all postnatal mothers who wanted the improved discharge assessment and counseling received it, regardless of their decision to enroll into the study. Study procedures, anticipated risks or discomfort, and potential benefits of the study were described to the potential participants. Informed verbal consent was undertaken with each participant upon completion of the discharge assessment and counseling. Confidentiality was maintained to the maximum standard during all stages of study implementation.

## Consent for publication

Not Applicable

## Data availability

The de-identified data that support the findings of this study are available from the corresponding author, upon reasonable request.

## Authors’ contributions

GB, DB, KW, DT, LA, WW, AH, LN, and AW contributed to the conceptualization of the study. GB and AW analysed and interpreted the data. GB drafted the manuscript. DB, KW, DT, LA, WW, PS, AH, LN, and AW contributed and guided the analysis and writing of the manuscript. All authors have read and approved the final manuscript.

## Acknowledgements

The authors express deepest gratitude to the study participants. Special thanks to field data collectors and supervisors. We would like also to thank the Ministry of Health of Ethiopia and Oromia Regional Health Bureau for their contribution during proposal development and for writing support letters to conduct the study.

## Additional files

Please find the additional files attached.

